# Plasma Metabolic Profiling Analysis of Heart Failure with Preserved Ejection Fraction Patients Based on UHPLC-MS/MS

**DOI:** 10.1101/2024.01.23.24301645

**Authors:** Muyashaer Abudurexiti, Tuersunjiang Naman, Duan dong qin, Salamaiti Aimaier, Ailiman Mahemuti

**Affiliations:** Department of Heart Failure, First Affiliated Hospital of Xinjiang Medical University, Xinjiang, China

**Keywords:** Heart failure, plasma metabolites, metabolomics, UHPLC-MS/MS, Tryptophan metabolism

## Abstract

**Background:** Heart failure with preserved ejection fraction (HFpEF) is a complex disease characterized by metabolic disturbances that can have various pathophysiological implications.

**Methods:** Data were collected from a prospective cohort consisting of 30 HFpEF participants and 30 healthy controls, matched by gender and age. untargeted metabolomic profiling UHPLC-MS/MS was performed on Venous blood. The analyses aimed to reveal the differences in plasma metabolic characteristics between HFpEF participants and healthy controls using untargeted metabolomics. Initial steps included Principal Components Analysis (PCA), Partial Least Squares-Discriminant Analysis (PLS-DA), and hierarchical clustering analysis to detect differing compounds groups. Subsequently, Receiver Operating Characteristic (ROC) curve analysis and pathway analysis of the different metabolites. Finally, we performed pathway enrichment analysis to identify significantly dysregulated processes.

**Results:** A total of 124 significantly different compounds were selected based on criteria of Variable Importance in Projection (VIP) > 1.0, Fold Change (FC) > 1.2 or FC < 0.833, and P-value < 0.05. Compared to healthy control group, the most altered metabolites in HFpEF were lipids and lipid-like molecules. KEGG enrichment and pathway impact analysis highlight the enrichment of these differentially expressed metabolites primarily in tryptophan metabolism. Hierarchical clustering results, showed varying compound levels between the two groups. ROC curve results, revealed Cytosine and 1,2-dihydroxyheptadec-16-yn-4-yl having higher AUC values.

**Conclusions:** Significant differences were observed in plasma metabolic profiles between HFpEF patients and healthy subjects using UHPLC-MS/MS. Cytosine appeared a potential biomarker. Tryptophan metabolism may be a novel metabolic pathway of HFpEF.

## 1. Introduction

Heart failure (HF) represents a complex clinical syndrome, and is often the end-stage of various cardiovascular diseases. Heart failure with preserved ejection fraction (HFpEF) is a subtype of HF that affects up to half of the estimated 65 million patients with HF worldwide [1]. However, its pathophysiology remains poorly understood. The diagnosis of HFpEF relies primarily on symptom evaluation, supplemented by physical examination, instrumental examination and the use of molecular biomarkers such as natriuretic peptides [2]. Nevertheless, most existing biomarker studies for HFpEF diagnosis exhibit a high risk of bias, limiting their clinical applicability and reproducibility [3].

Metabolic alterations have been implicated in the development and progression of various cardiovascular diseases, including heart failure [4–6]. Nonetheless, the majority of these studies utilized animal models, leading to a paucity of data on human HFpEF metabolism. Concurrently, has been a resurgence of interest in metabolic impairment as a potential contributing factor to the onset and progression of HFpEF [1,7–9]. Therefore, investigating the plasma metabolic profile of HFpEF patients could provide valuable insights into the underlying mechanisms and potentially identify novel biomarkers for early diagnosis and targeted therapies [10].

In recent years, advances in analytical techniques, such as ultra-high-performance liquid chromatography coupled with tandem mass spectrometry (UHPLC-MS/MS), have facilitated the comprehensive and precise profiling of metabolites in biological samples. High sensitivity, selectivity, and throughput, are provided by UHPLC-MS/MS, rendering it an ideal platform for metabolomic analysis [11,12]. This study, employed untargeted metabolomics to compare metabolite expression profiles between HFpEF patients and healthy controls, to uncover new insights and potential therapeutic targets for HFpEF.

## 2. Materials and methods

### 2.1. Participants and clinical samples collection

HFpEF Study Population: Samples were collected from 30 HFpEF patients and 30 healthy controls at the First Affiliated Hospital of Xinjiang Medical University between March 1, 2023, and July 31, 2023. participants, aged 18 to 85 years, provided informed consent.

Diagnosis Criteria: patients were diagnosed with HFpEF based on consensus criteria [4,5,13]: which include symptoms and signs of exertional dyspnea (New York Heart Association class II or III), HF with left ventricular ejection fraction (LVEF) ≥ 50%, and at least two of the following: (1) elevated NT-proBNP (N-terminal pro-B-type natriuretic peptide) ≥125 pg/mL; (2) structural heart disease or diastolic dysfunction on echocardiography; (3) E/e’≥9;

Exclusion Criteria: Excluded were Patients with a history of congenital heart disease, LVEF < 40%, HF with mid-range EF (40–50%), hypertrophic cardiomyopathy, cardiac transplantation, constrictive pericarditis, severe valvular disease, or infiltrative or restrictive cardiomyopathy [1,14];

Ethical Approval: This study received approval from the Ethics Committee of the First Affiliated Hospital of Xinjiang Medical University. informed consent was obtained from each participant.

Sample Collection: Overnight – fasting Venous blood samples were collected in the morning before breakfast. after centrifugation at 3000 rpm for 10 min, supernatant were collected and stored at−80 °C until analysis.

### 2.2. Plasma sample preparation

Sample Processing: Plasma samples, collected using EDTA tubes, were processed immediately. Each 100μL sample was resuspended in pre-chilled 80% methanol and vortexed thoroughly. Following 5 min incubation on ice and 20 min centrifugation at 15,000 g at 4°C, supernatants were collected and diluted with LC-MS grade water to a final concentration 53% methanol.

LC-MS/MS Analysis: diluted samples underwent a further 20 min centrifugation at 15,000 g at 4°. The supernatants were then subjected to LC-MS/MS analysis.

Quality Control: quality control (QC) samples, comprising equal volumes mixtures of experimental samples, were prepared to monitor chromatography-mass spectrometry system balance, system stability, and instrument status throughout the experiment. blank samples were also included to eliminate background ions.

### 2.3. UHPLC-MS/MS analysis

UHPLC-MS/MS analyses was conducted using the Vanquish UHPLC system (ThermoFisher, Germany) coupled with an Orbitrap Q Exactive^TM^ HF or Orbitrap Q Exactive^TM^HF-X mass spectrometer (Thermo Fisher, Germany) at Novogene Co., Ltd. (Beijing, China). Samples were injected onto the Hypersil Gold column (100×2.1 mm, 1.9μm) at a flow rate of 0.2 mL/min over a 12min linear gradient. positive ion mode eluents included 0.1% formic acid in water (eluent A) and Methanol (eluent B), while negative ion mode eluents consisted of 5 mM ammonium acetate, (pH 9.0, eluent A) and methanol (eluent B).The elution profile was as follows: 1.5 min with 2%B;3 min with 2-85% B;10 min with 85-100% B; 10 min with 100-2% B and 12 min with 2% B. The Q Exactive^TM^ HF mass spectrometer operated under the following conditions: positive/negative ion mode,3.5 kV spray voltage, 320°C capillary temperature, 350°C Aux gas heater temperature, 10 L/min aux gas flow rate, 35 psi sheath gas flow rate, and an S-lens RF level of 60.

### 2.4. Data processing and metabolite identification

raw data from plasma samples were obtained via UHPLC-MS/MS and processed using Compound Discoverer 3.1 (CD3.1, Thermo Fisher) for peak picking, alignment, and metabolite quantitation key parameters included a 0.2 min retention time tolerance, 5ppm mass tolerance, a signal/noise ratio of 3, 30% signal intensity tolerance, and a minimum intensity threshold. Subsequently, peak intensities were normalized relative to total spectral intensity. predicting molecular formulas was based on molecular ion peaks, fragment ions, And additive ions. Peak matching against the mzCloud (https://www.mzcloud.org/), mzVault and Mass List database was carried out to obtain relative and accurate qualitative data. Statistical analyses were conducted using R (R-3.4.3), CentOS (CentOS release 6.6), and Python (Python 2.7.6), For normally distributed data, standardization was achieved using the formula: raw quantitation value of the sample / (total metabolite quantitation value of the sample/total QC1 sample metabolite quantitation value). compounds with a CV over 30% in QC samples were excluded, enabling metabolite identification and quantification.

### 2.5. Data Analysis

Metabolite annotation in plasma samples utilized the KEGG (https://www.genome.jp/kegg/pathway.html), LIPIDMaps (http://www.lipidmaps.org/) and HMDB (https://hmdb.ca/metabolites) database. Using metaX [15] Partial least squares discriminant analysis (PLS-DA) and principal components analysis (PCA) were conducted. A univariate regression (t-test) determined significant differences (P-value). Metabolites meeting criteria of VIP >1 and P-value< 0.05 and fold change≥2 or FC≤0.5 were classified as differentially expressed. Volcano plots generated by ggplot2 in R, facilitated the selection of metabolites based on log2(FC) and −log10(P-value).

## 3. Results

### 3.1. Baseline features of participants

Table 1 compares the basic characteristics of HFpEF and HC groups. The groups were matched for age and gender. Compared with the two groups, HFpEF subjects presented significantly higher levels of IL-6, LP(b), NT-ProBNP, Cr and lower levels of TC,LDL,HDL compared to the HC group, with notable incidences of hypertension (70%, *p*=0.004), CAD (56.7%, *p*<0.001), DM (46.7%, *p* <0.001). However, lower levels of TC and LDL, HFpEF patients might be attributable to lipid-lowering treatments. Other parameters like CRP, TG, LP(a), E/è, LVED and SAA showed no significant differences between groups (P>0.05).

**Table 1.**
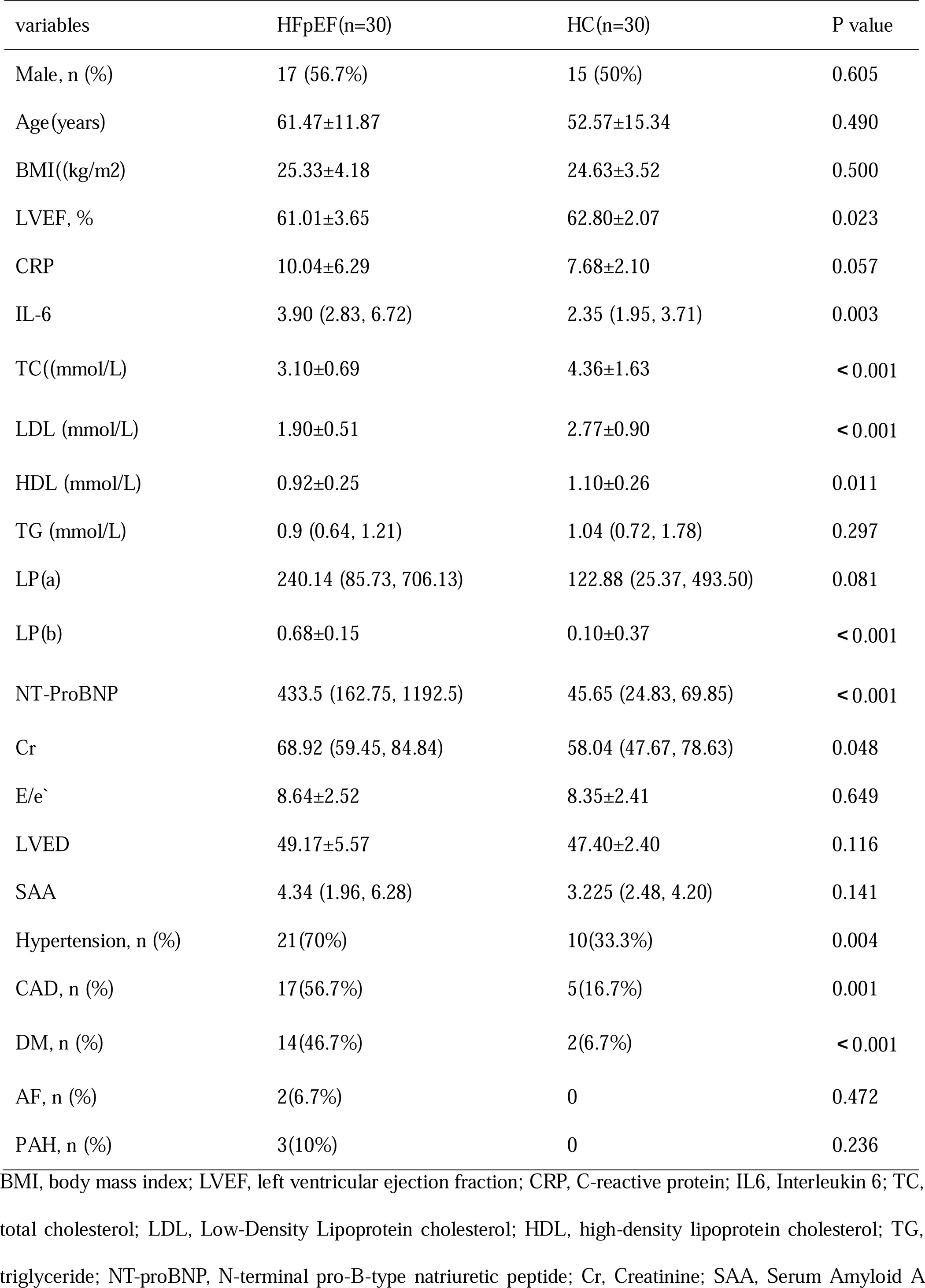
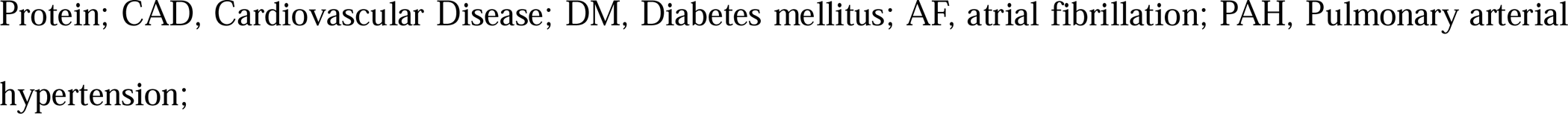
Basic characteristics of the participants.

### 3.2. Quality control of untargeted metabolic profiling

Considering the influence of exogenous factors, on the metabolome, ensuring instrumental stability and normal signal response in metabolite detection was critical QC was performed using Pearson correlation analysis between QC samples and principal component analysis (PCA). Pearson correlation coefficients (R2) among QC samples were close to 1 in both ion modes (Figure 1A, B), indicating high stability and data quality. tightly clustering in PCA, of QC samples further confirmed the stability and reproducibility of instrumental analysis (Figure1C, D).

**Fig. 1.**
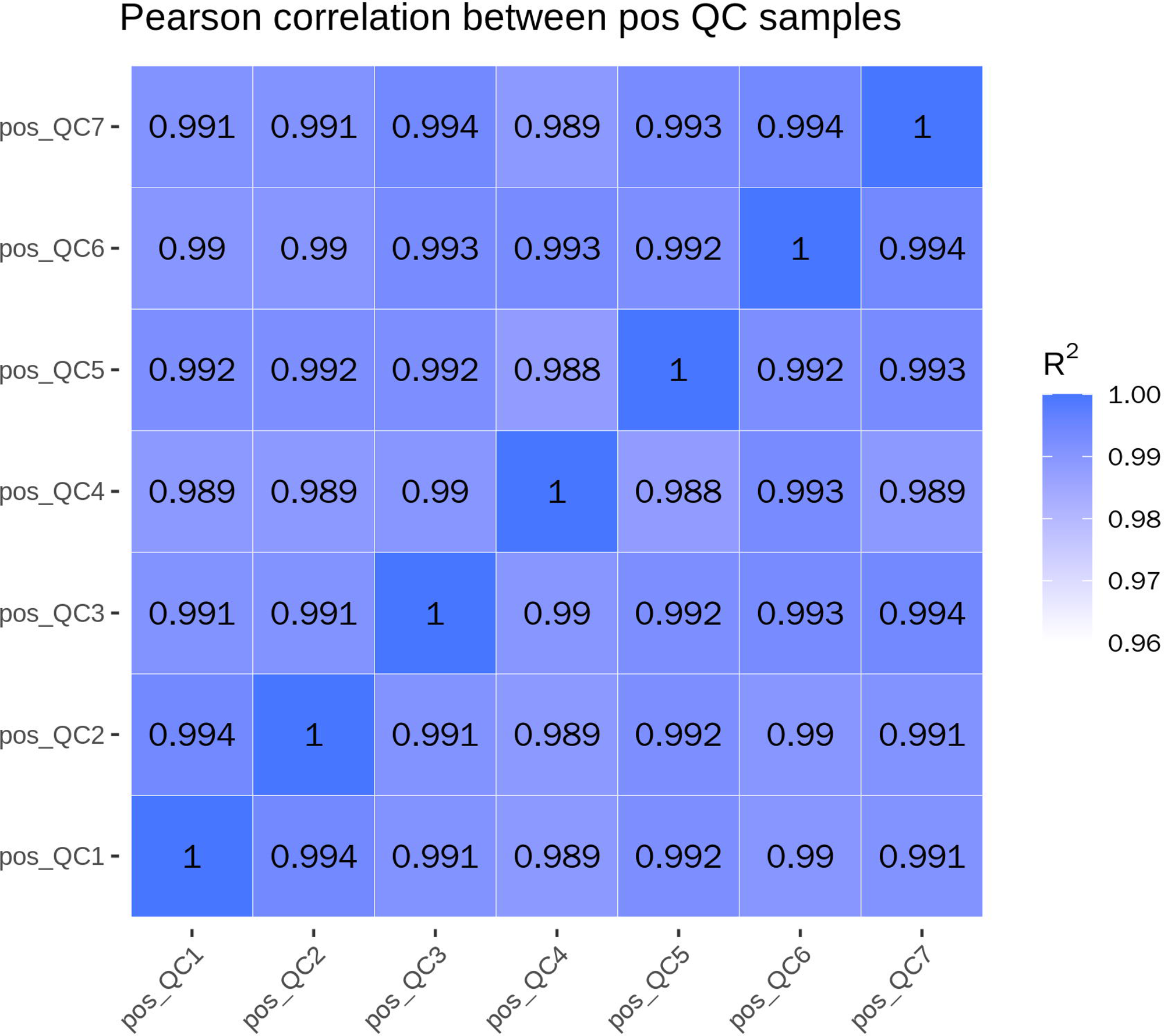
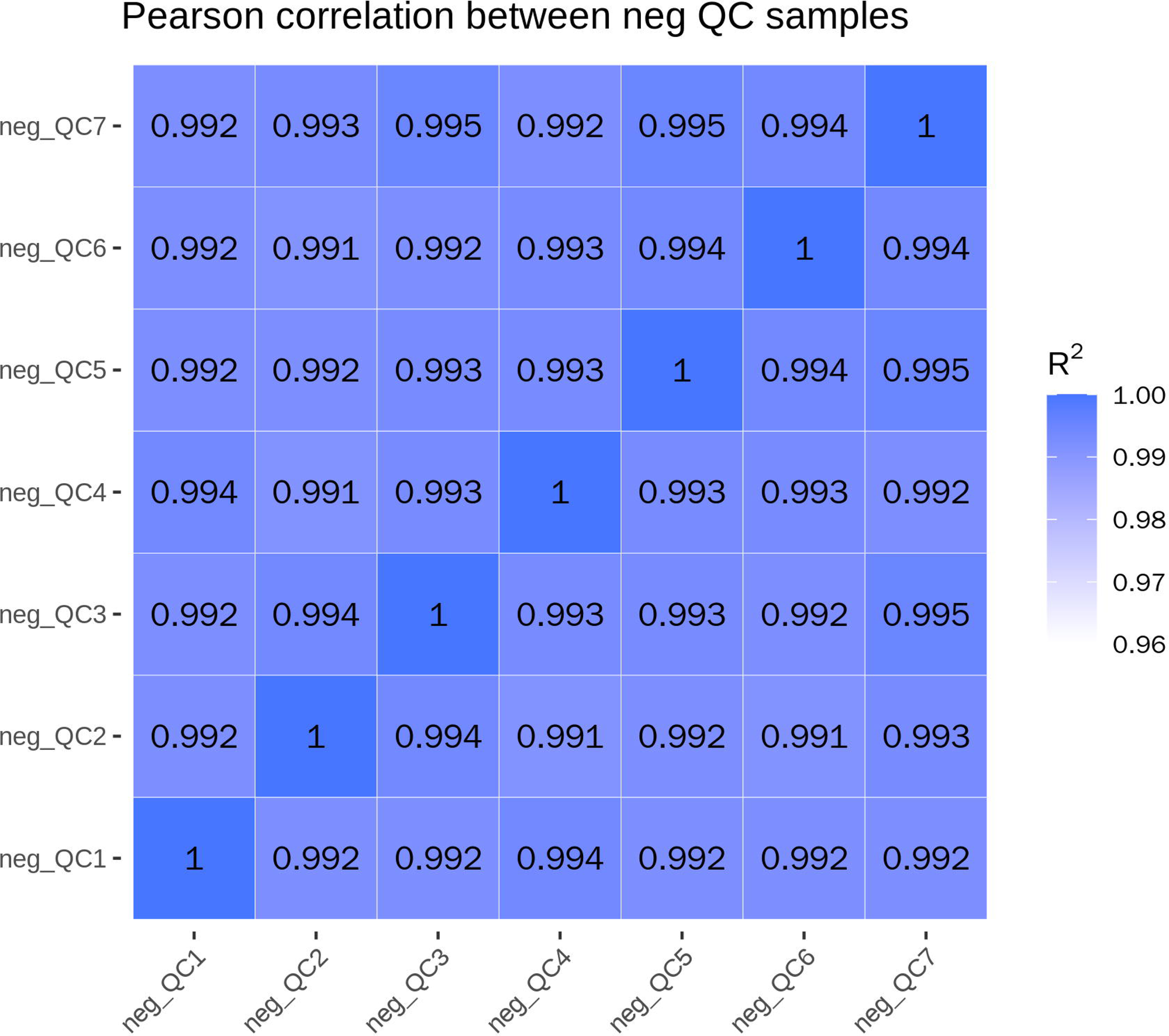
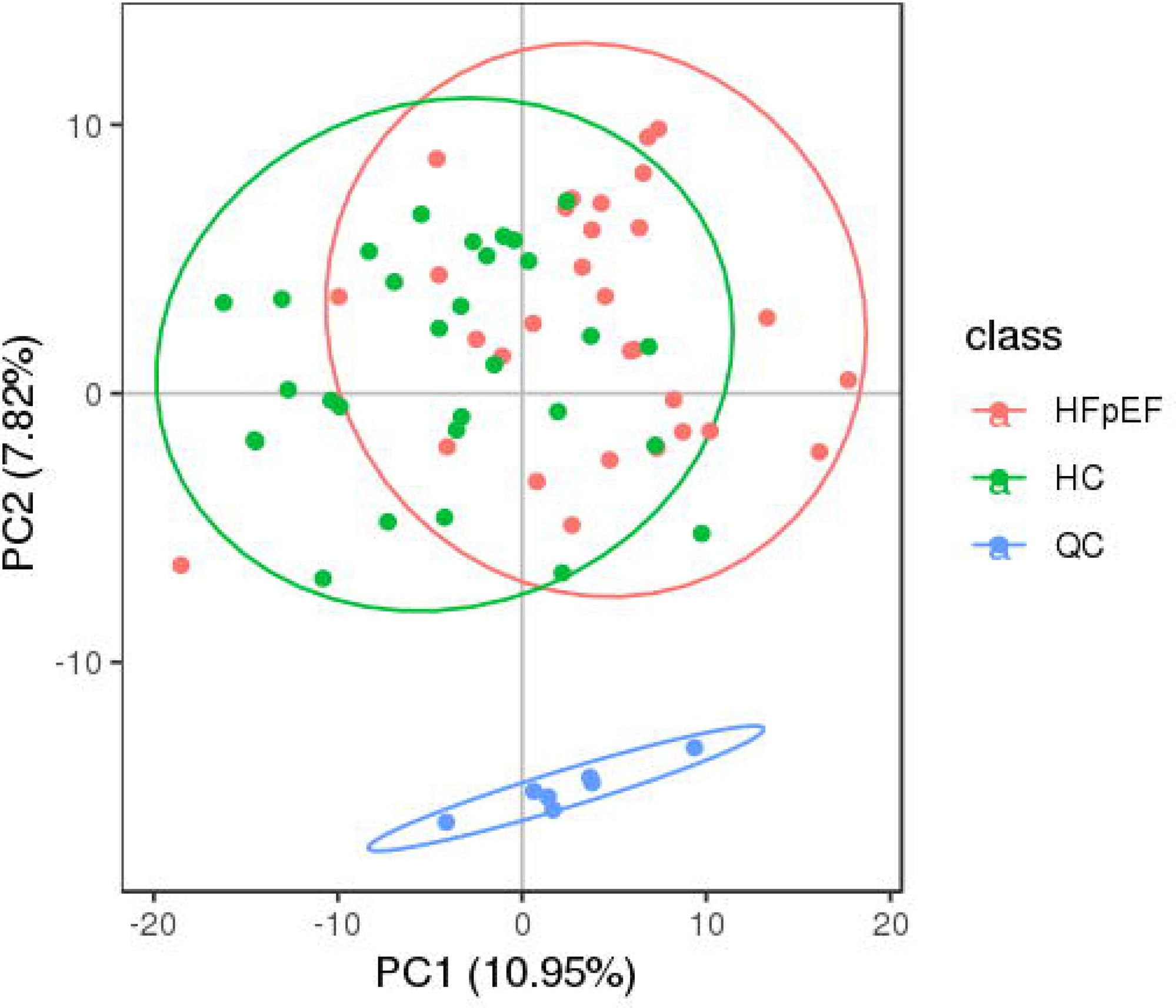
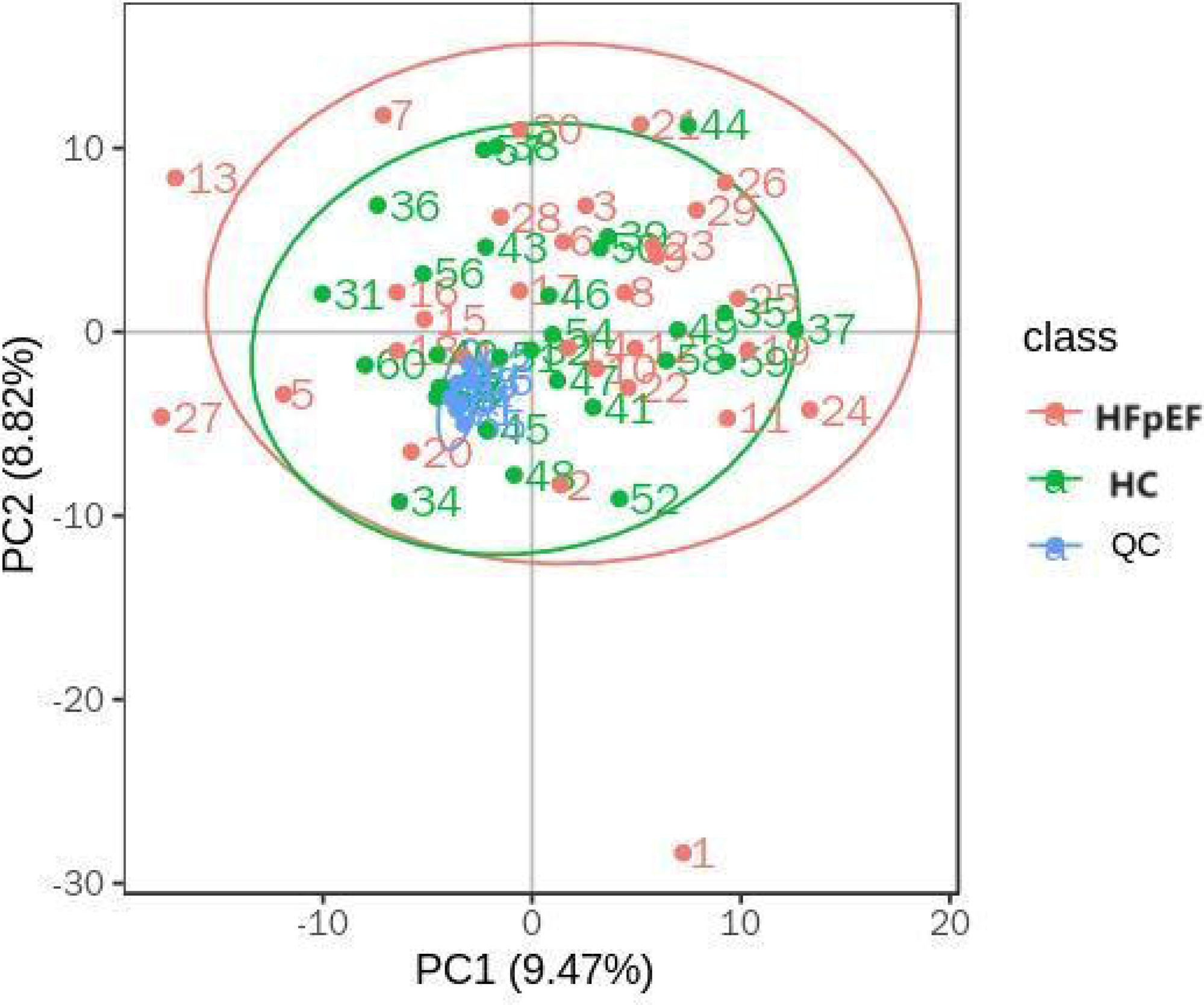
The quality control of untargeted metabolomic profiling. (A, B) Pearson correlation analysis between QC samples: the coefficient (R2) values were both nearly 1 under the positive (A) or negative (B) polarity modes. (C,D) PCA analysis:QCs tightly aggregated into a cluster in the positive (C) and negative (D) ion modes.

### 3.3. Metabolite pathways and classification annotations

Comparative analysis of metabolic signatures in untargeted metabolomics between HFpEF and HC groups under ESI+ and ESI− modes. identified 993 Metabolites MS1 and MS2 data were compared with compounds from mzCloud, mzVault, Mass List, KEGG, Human Metabolome Database (HMDB) and LIPID Maps databases for metabolite identification. A total of 124 metabolites were significantly differentially expressed in the HFpEF group.

### 3.4. Differential Metabolites Identification

Investigation of alterations in various metabolites within the HFpEF group, necessitated the application of multivariate statistical methodologies, specifically PCA and PLS-DA to elucidate the relationship between biological features and metabolomics. Unsupervised PCA, a conventional approach in pattern recognition, was employed for scrutinizing the distribution oof the HFpEF group and the removal of outlier data. PCA revealed the distinctiveness of the two groups based on PC1 and PC. Concurrently, disparities in metabolite profiles between the HFpEF and HC groups were observed (Figure 2A). Furthermore, supervised PLS-DA multivariate analysis corroborated significant dissimilarities in both groups, unveiling distinct clustering of the HFpEF and HC groups (Figures 2B and 2C). Additionally, under positive/ and negative-ion modes, classification parameters (R2Y) were determined to be 0.84 and 0.82, respectively, while Q2Y data were 0.48 and –0.08, respectively. These results denote a favorable model fit and predictive performance.

**Fig. 2.**
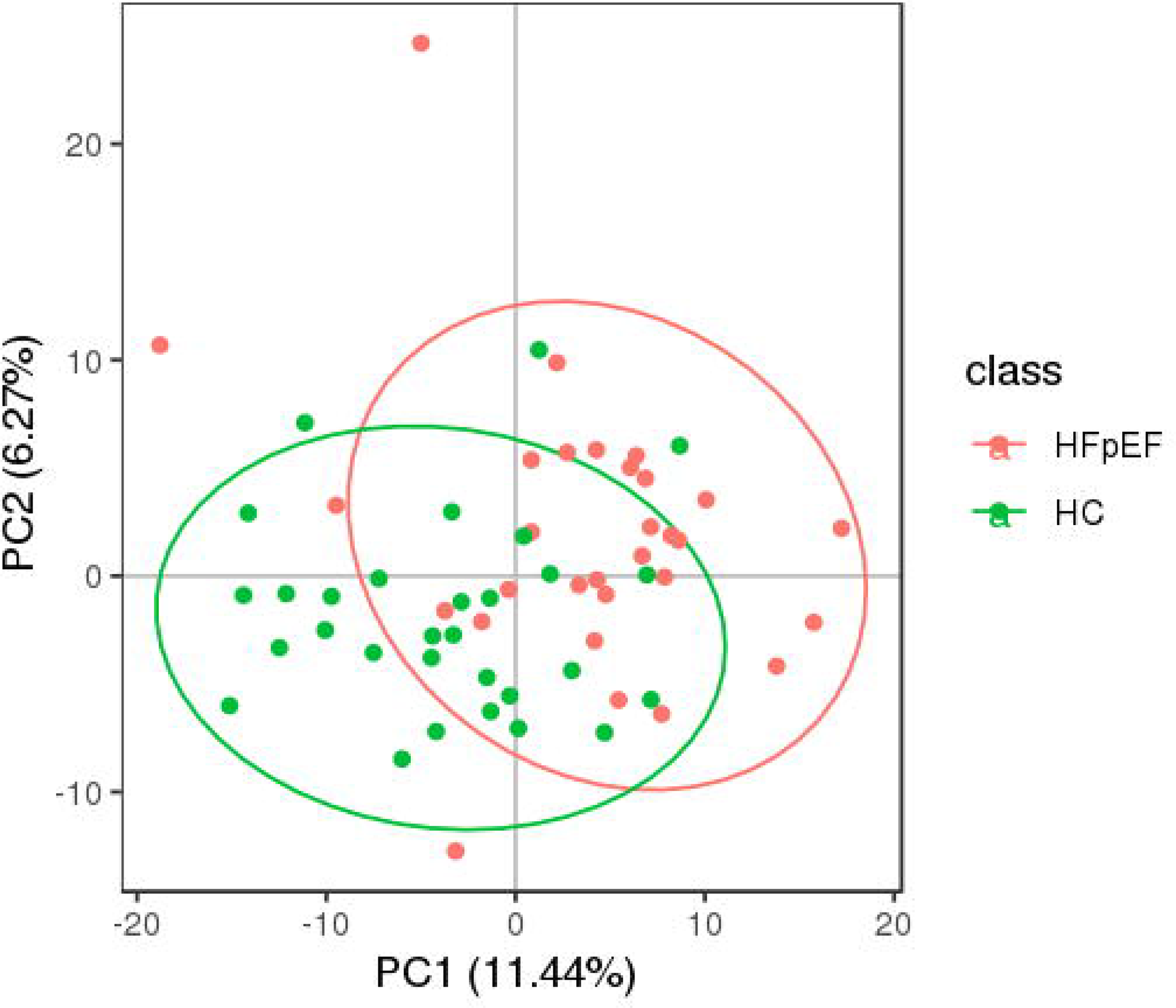
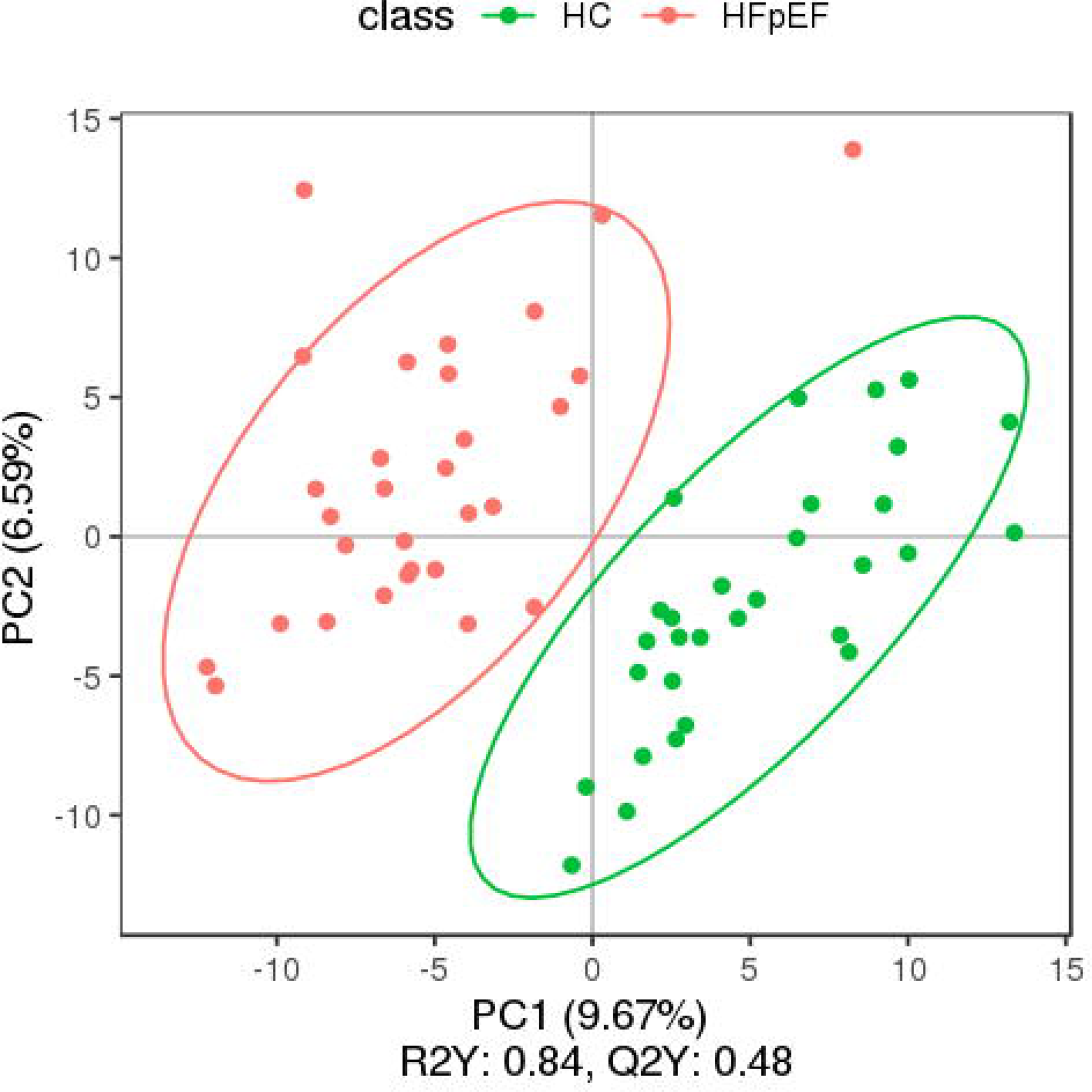
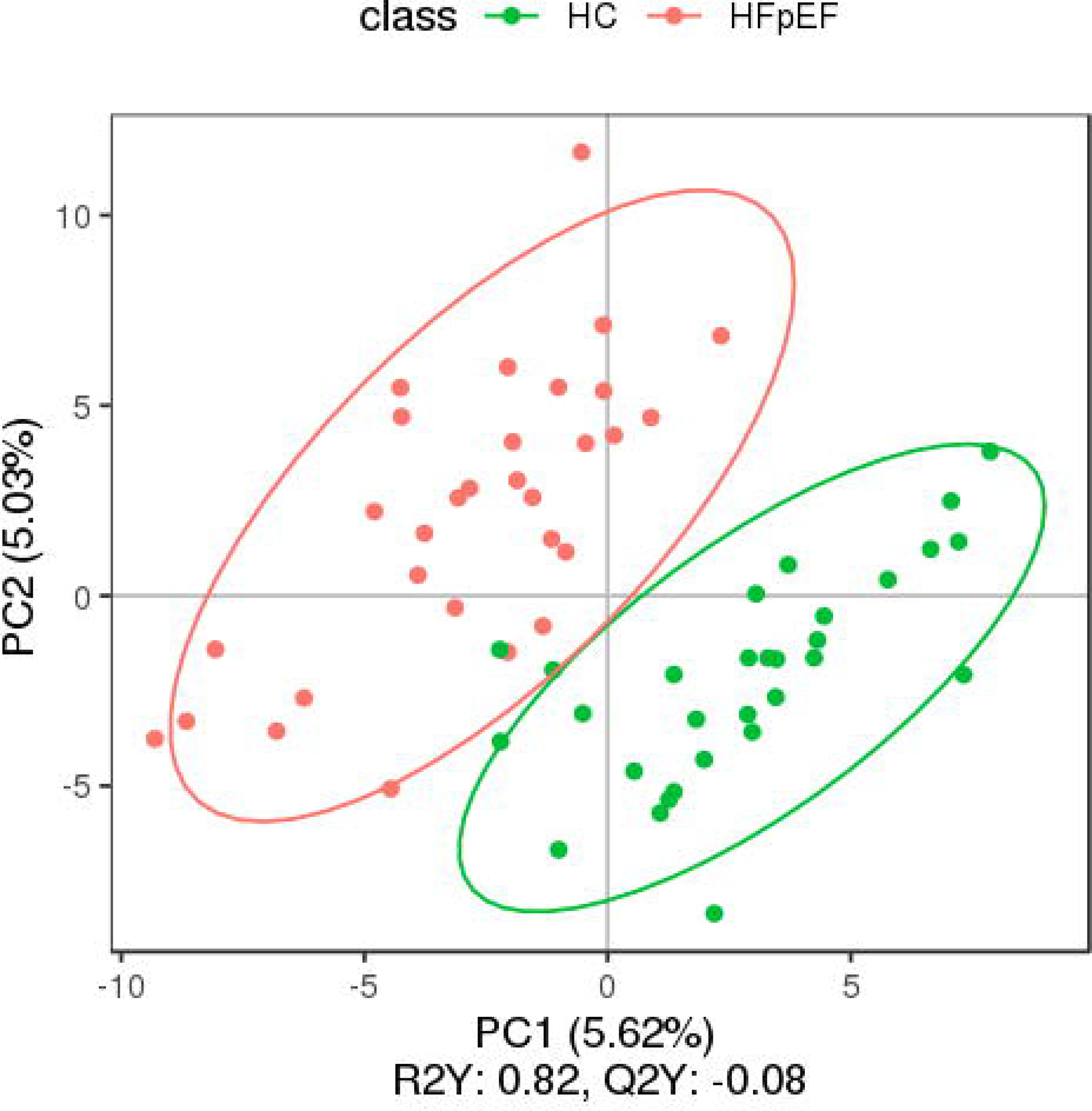
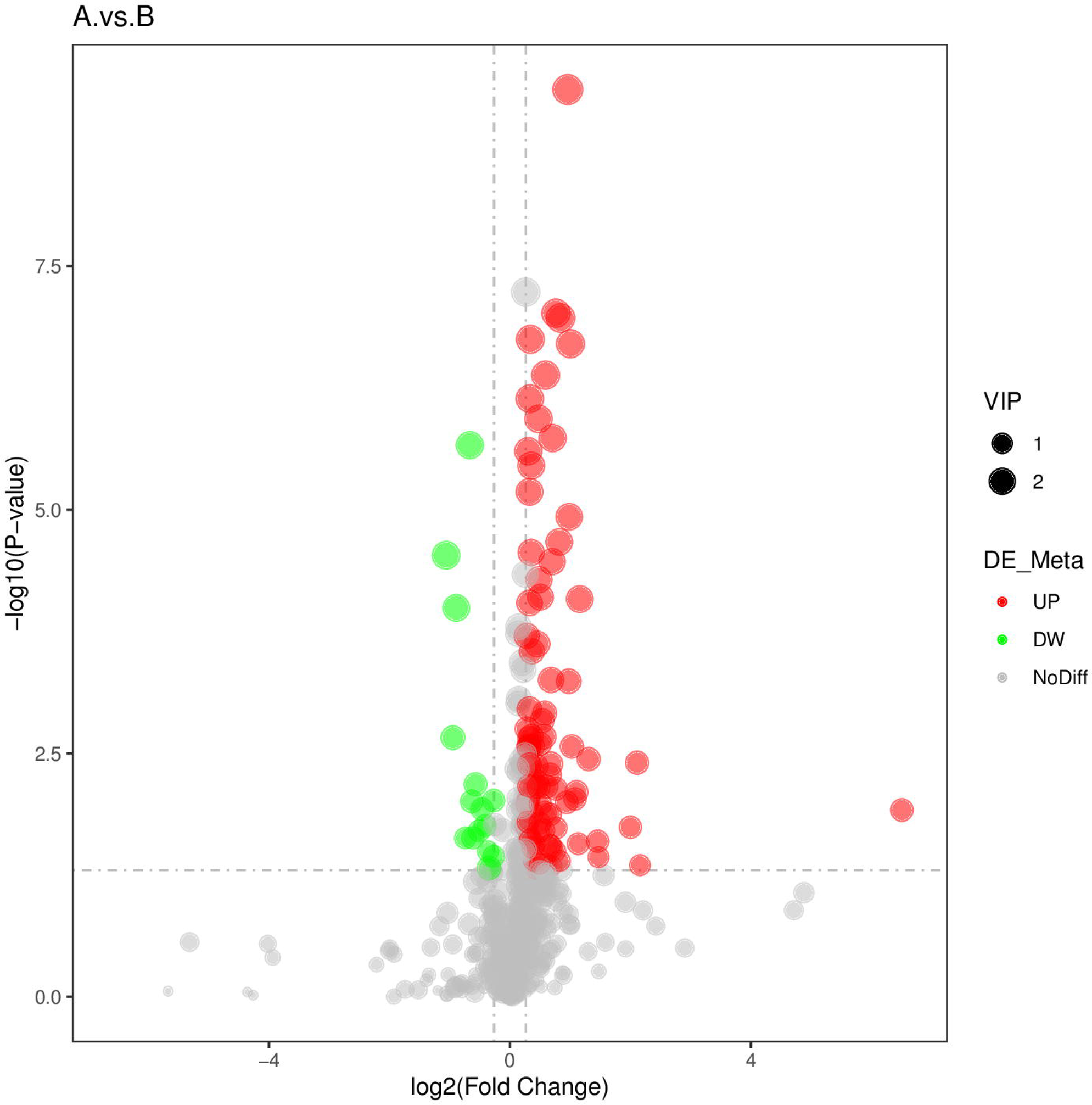
(A) PCA analysis comparing the two groups. (B and C) PLS-D analysis of differential metabolites in HFpEF and HC groups. (B) PLS-DA for fecal samples in negative ion mode. (C) PLS-DA for fecal samples in positive ion mode. (D) Volcano plot illustrating differential metabolite expression between HFpEF and HC groups.

To elucidate the characteristics of plasma metabolites within the HFpEF group and identify metabolites confidently associated with HFpEF, distinctions under ESI+ and ESI-modes were made based on VIP (Variable importance in projection, VIP) > 1.0, FC > 1.2 or FC < 0.833 and a P-value< 0.05.Consequently, 993 differential compounds were identified from plasma samples, with 124 of them reaching statistical significance, among these, 87 metabolites exhibited upregulation, with fold changes reaching up to 2.77, and 15 metabolites displayed downregulation, with fold changes as low as 0.48. A volcano plot was generated to visualize the distribution of differential metabolites between the two groups (Figure2D). Additionally, a heat-map was constructed to depict the significantly distinct plasma metabolites in HFpEF versus HC groups (Figure 3A). Notably, it was discerned that the significantly upregulated metabolites in the HFpEF group comprised lipids and amino acids, whereas the significantly downregulated metabolites consisted of phosphatidylcholines (PCs) and phosphatidylethanolamine (PEs) (Table2);

**Fig. 3.**
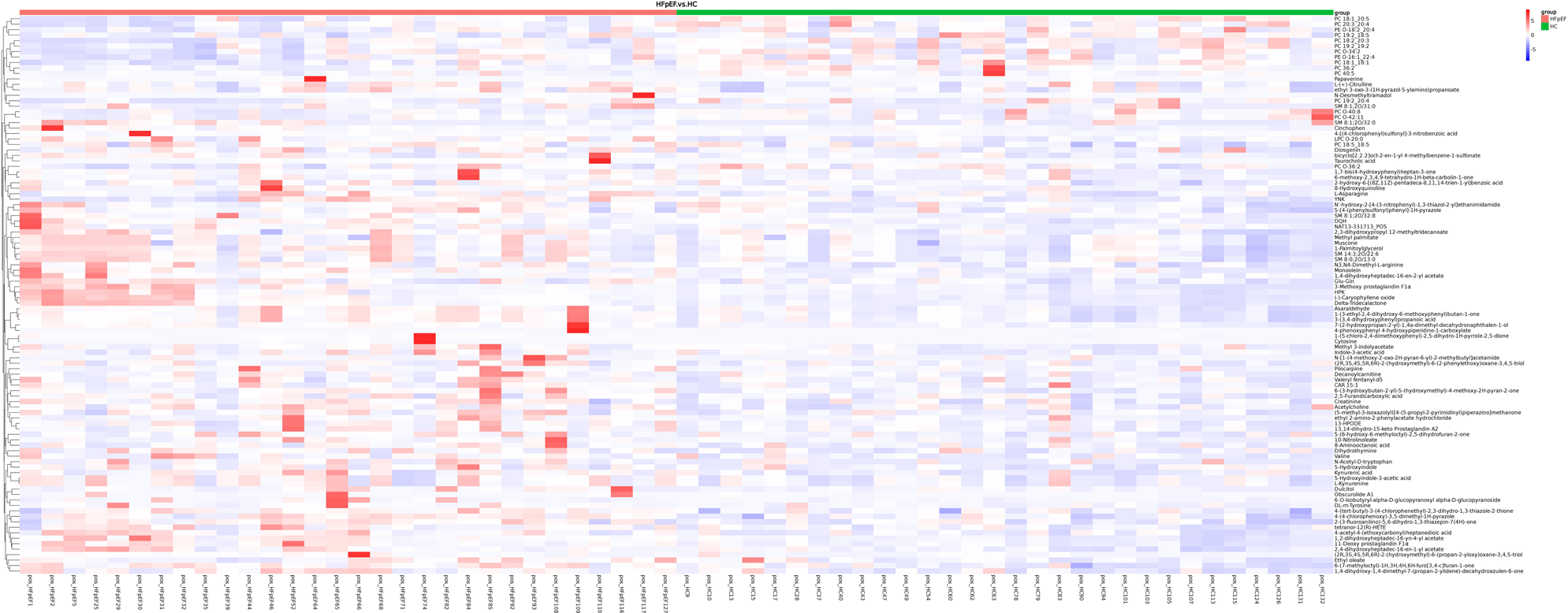
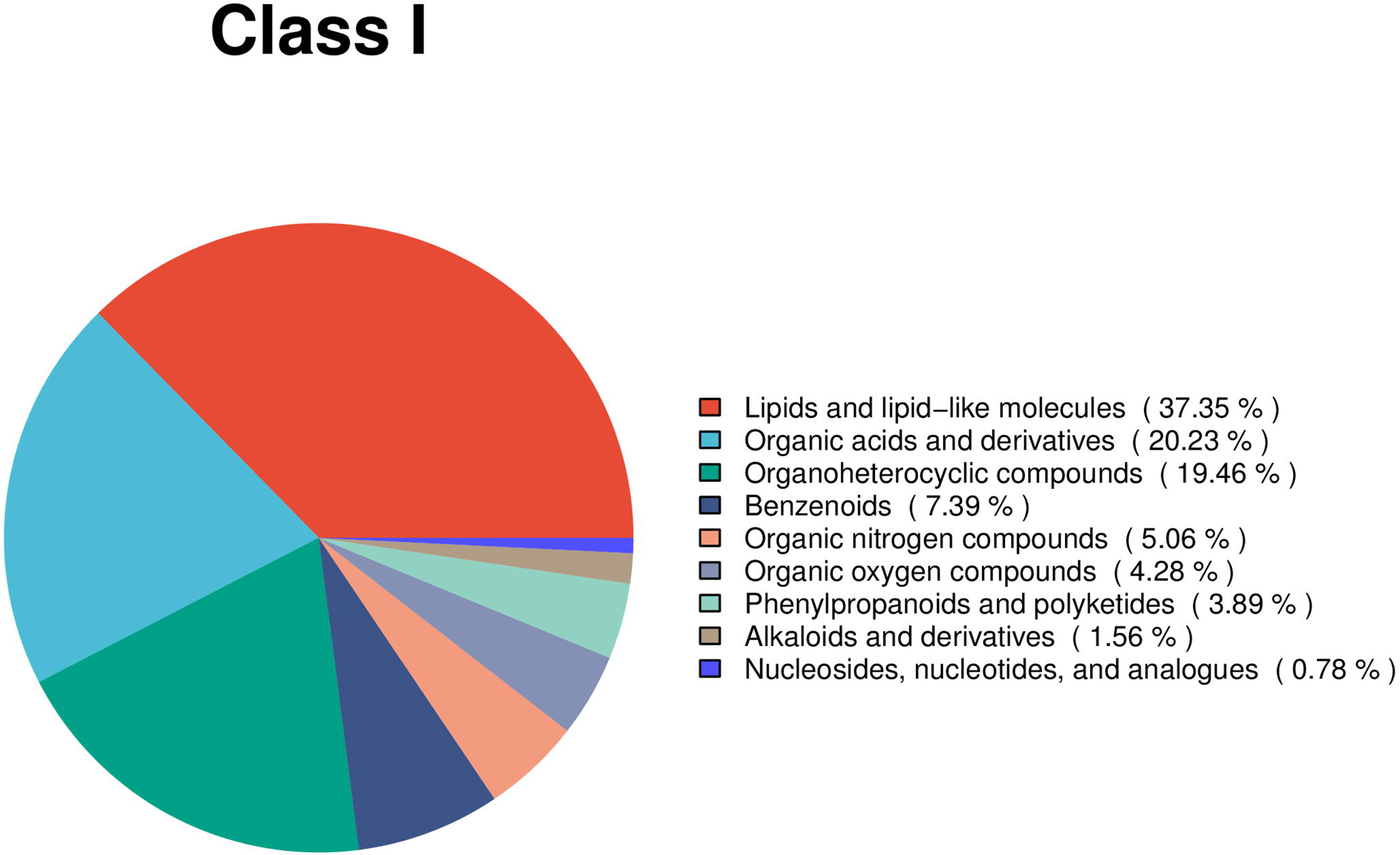
(A) Differential clustering analysis. (B) Classification of differential metabolites identified in HMDB database.

**Table 2.**
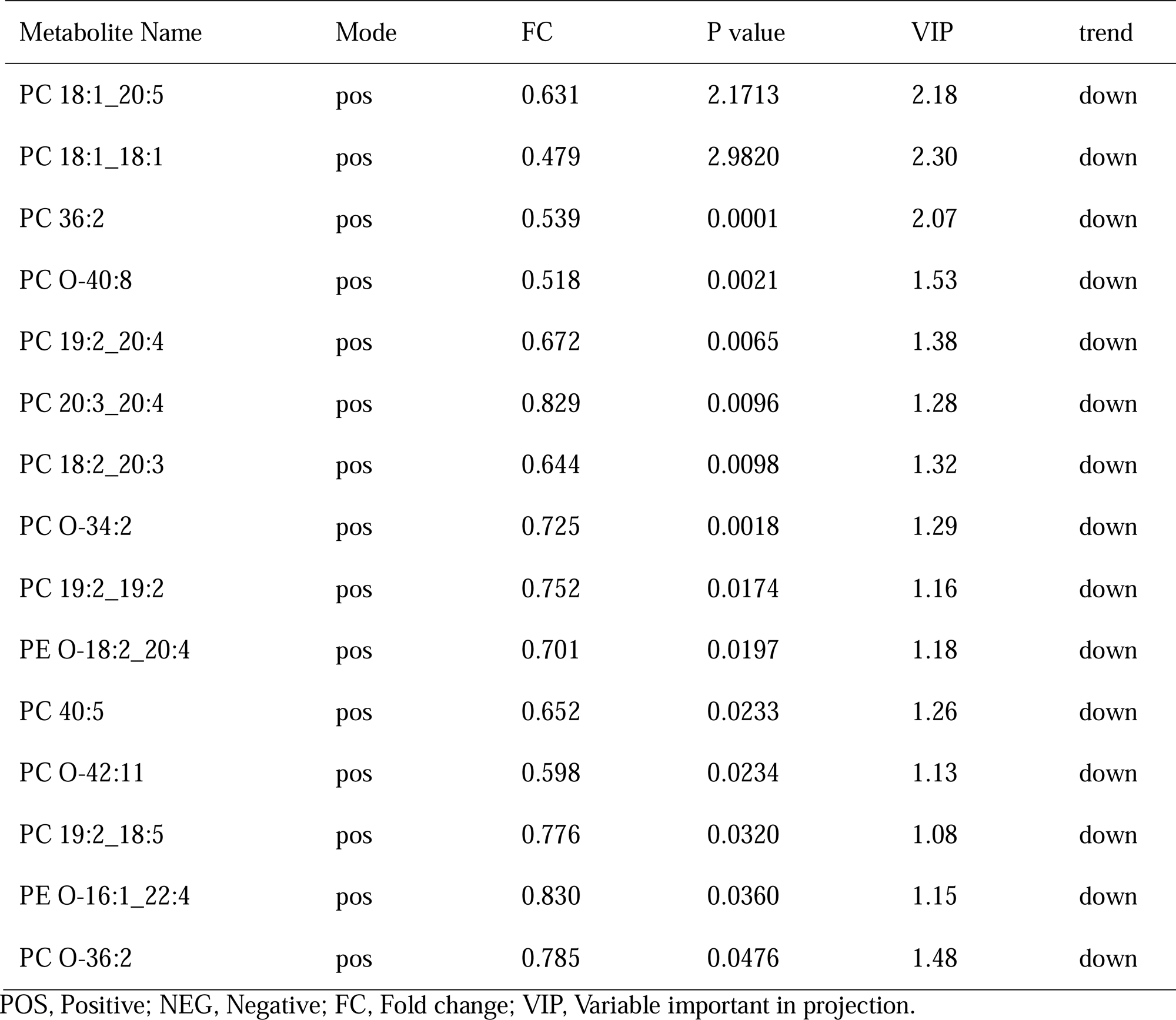
Downregulated metabolites identified from metabolomics profiling.

Subsequently, 32 putative metabolites that matched entries in the HMDB were verified. Among these metabolites, amino acids, peptides, and analogs (15.62%), and Fatty acid (FA) esters (9.38%) constituted the predominant chemical classes (Figure 3B).

### 3.5. Significance of differential metabolites in diagnosing HFpEF

Univariate ROC curves were generated for each metabolite to assess their diagnostic potential for HFpEF. In this investigation, it was ascertained that Cytosine, 1,2-dihydroxyheptadec-16-yn-4-yl acetate, PC 18:120:5, and PC 18:118:1 could potentially serve as biomarkers for HFpEF (Figure 4). These metabolites also exhibited the highest significance, with Cytosine and 1,2-dihydroxyheptadec-16-yn-4-yl acetate demonstrating superior Area Under the Curve (AUC) values.

**Fig. 4.**
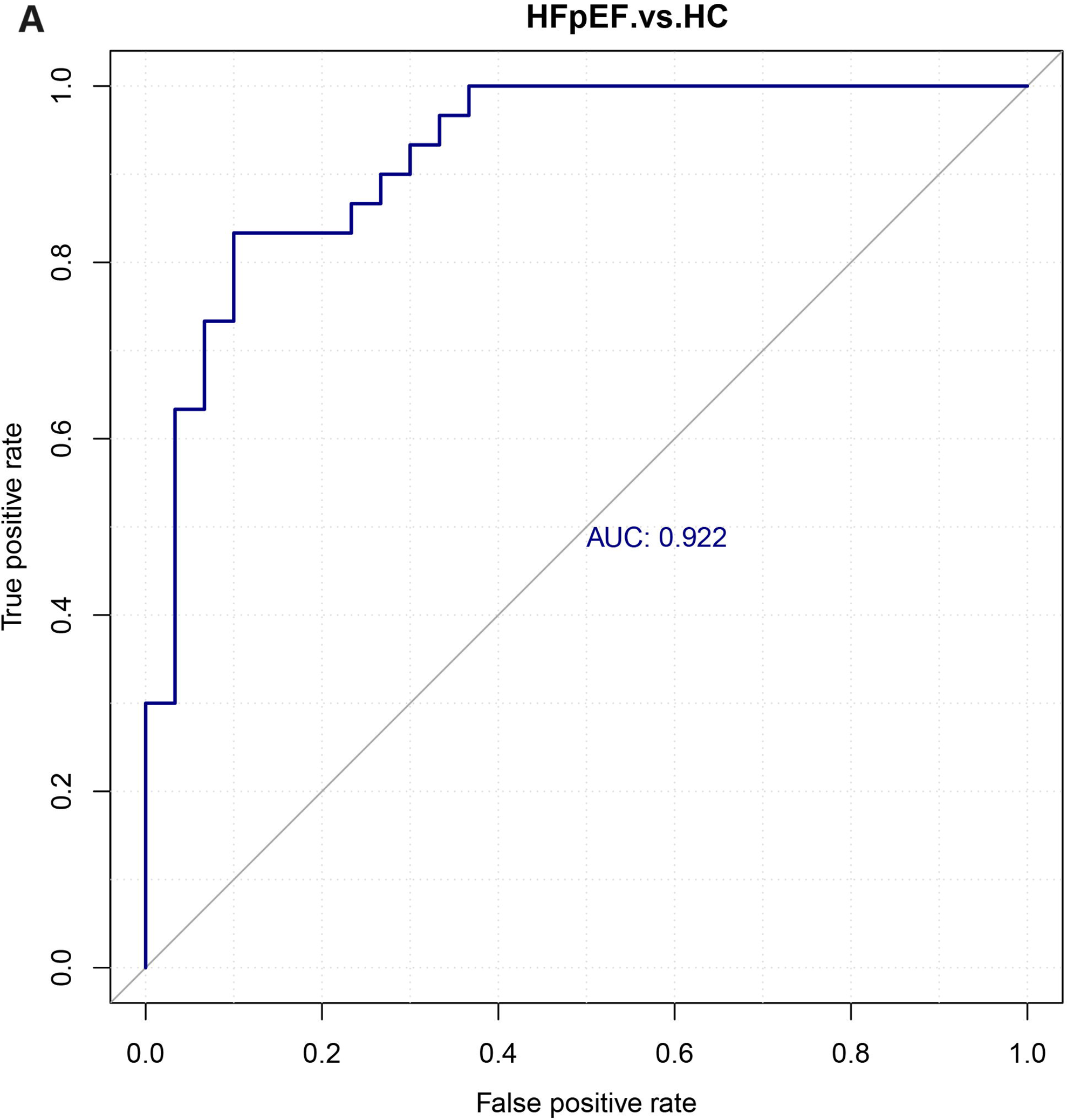
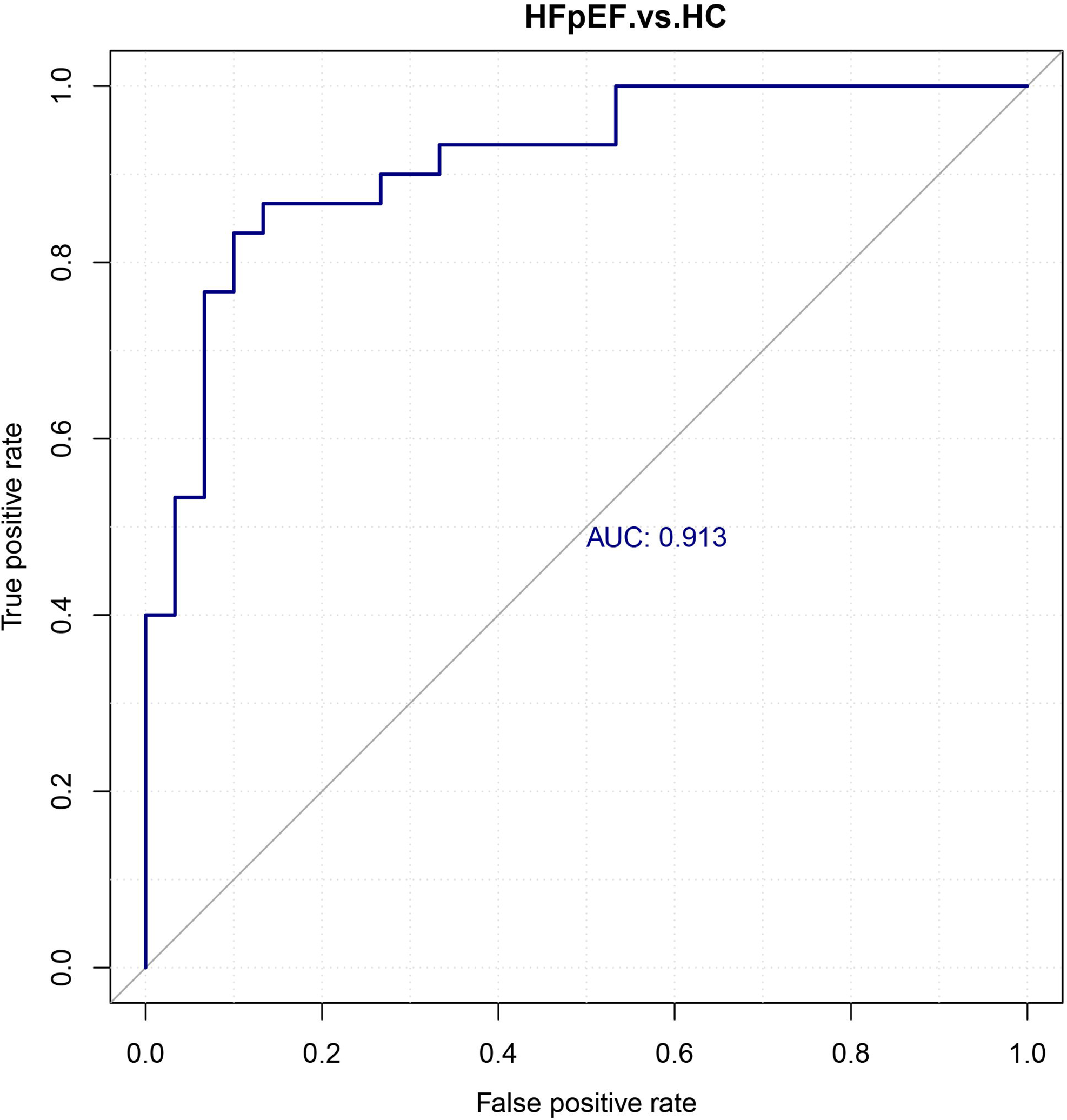
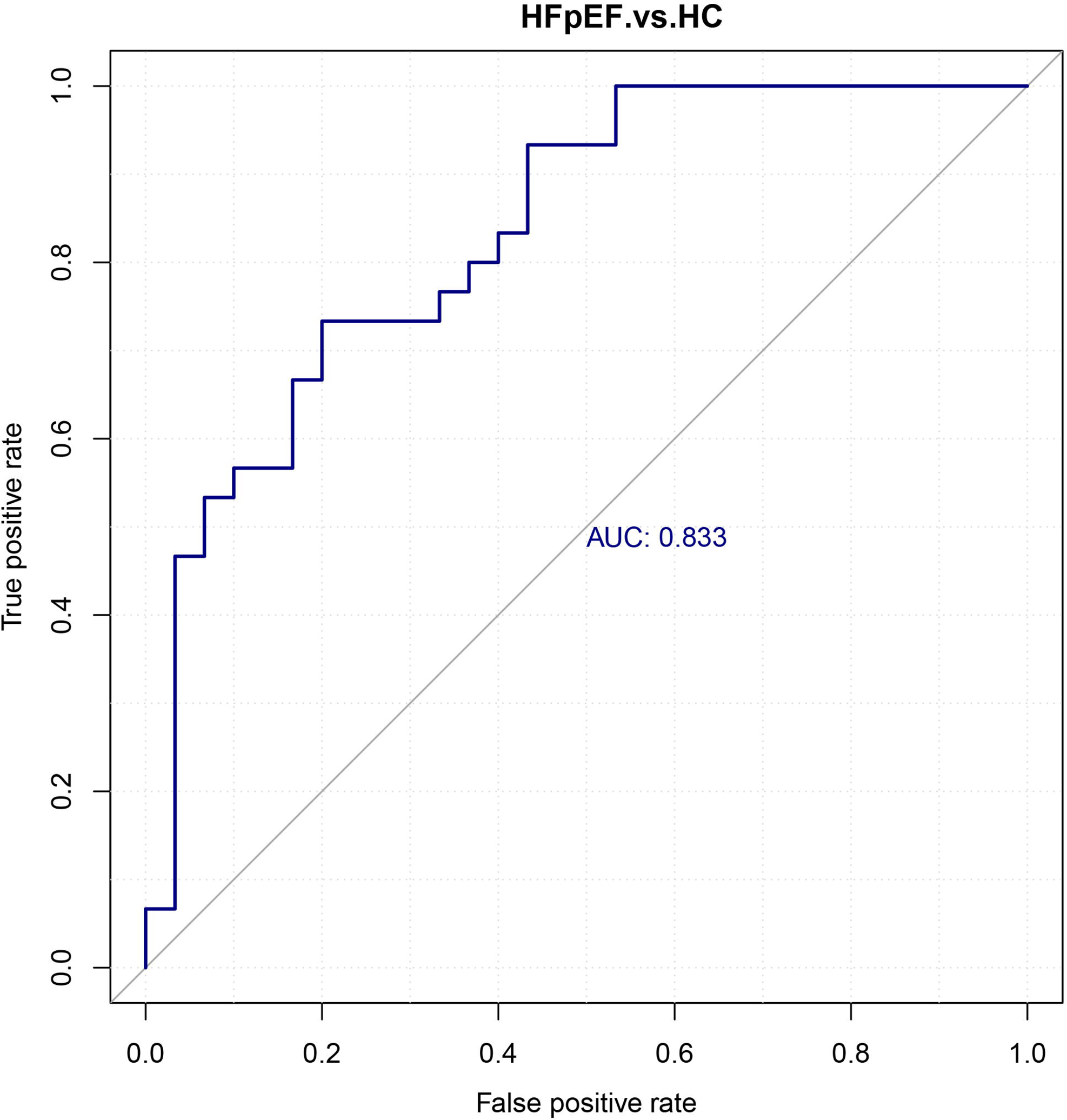
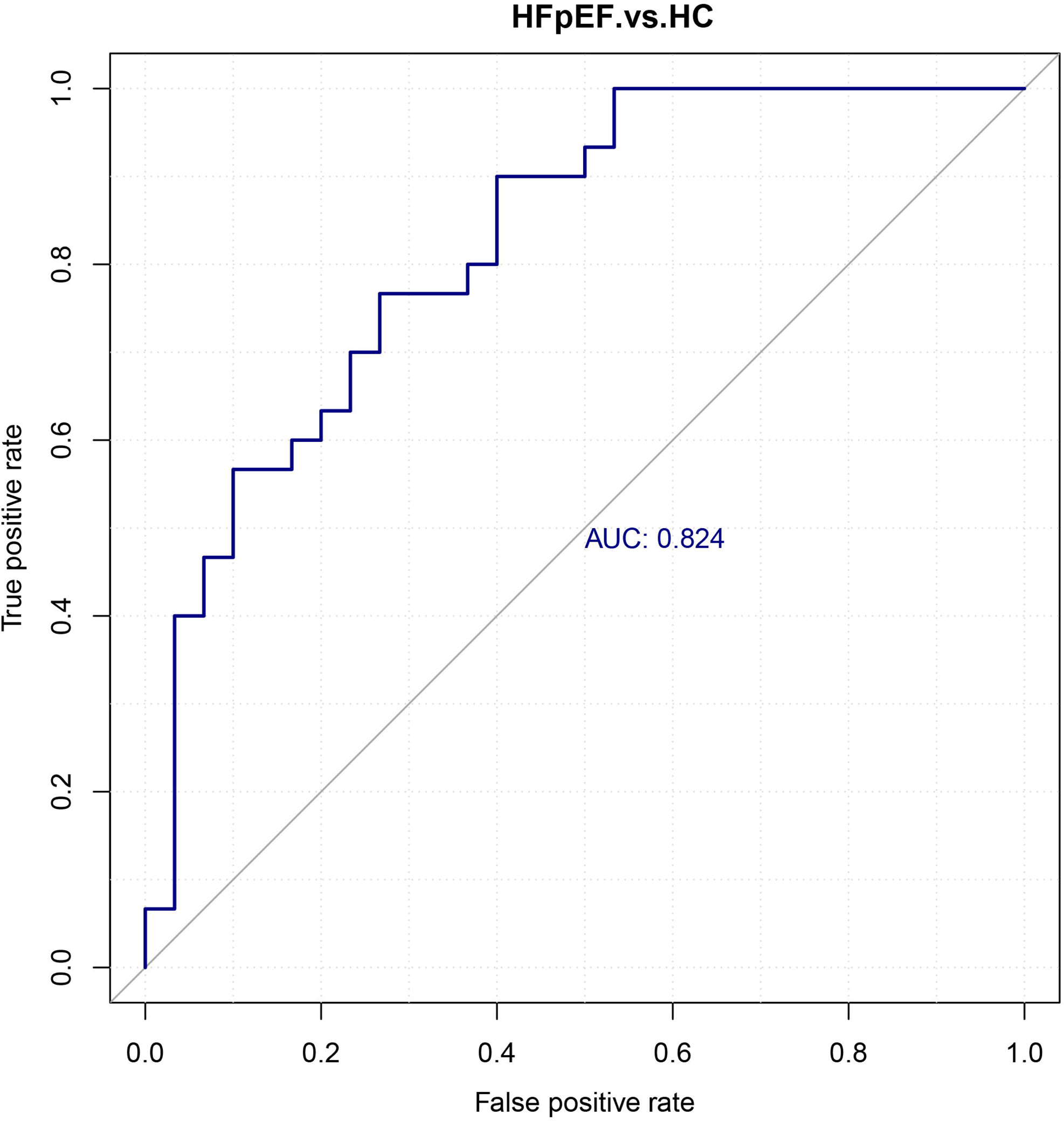
The receiver operating characteristic (ROC) analysis of different metabolites. A (Cytosine). (B) 1,2-dihydroxyheptadec-16-yn-4-yl acetate. (C) PC 18:1_20:5. (D) PC 18:1_18:1.

### 3.6. Pathway enrichment

Differential metabolites in both groups underwent pathway enrichment analysis based on the enrichment factor and P-value. This analysis revealed associations with several pathways, including Tryptophan metabolism, Metabolic pathways, Galactose metabolism, Primary bile acid biosynthesis, Steroid biosynthesis, Endocrine and other factor-regulated calcium reabsorption. Moreover, three differential metabolites were specifically associated with Tryptophan metabolism. The KEGG enrichment plot is illustrated in Figure 5. These differential metabolites showed significant enrichment in metabolic pathways such as Tryptophan metabolism, Tuberculosis and Endocrine and other factor-regulated calcium reabsorption.

**Fig. 5.**
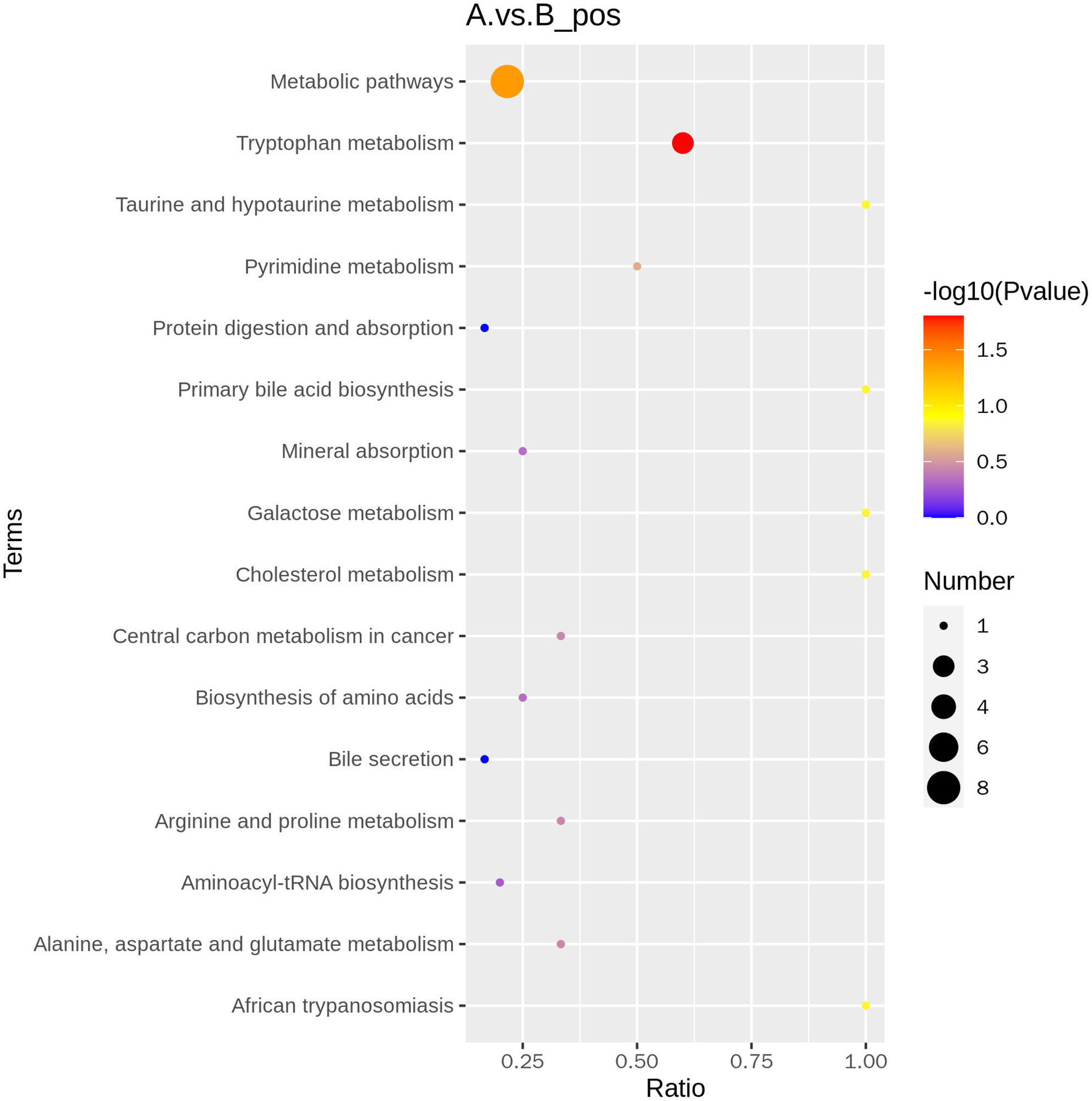
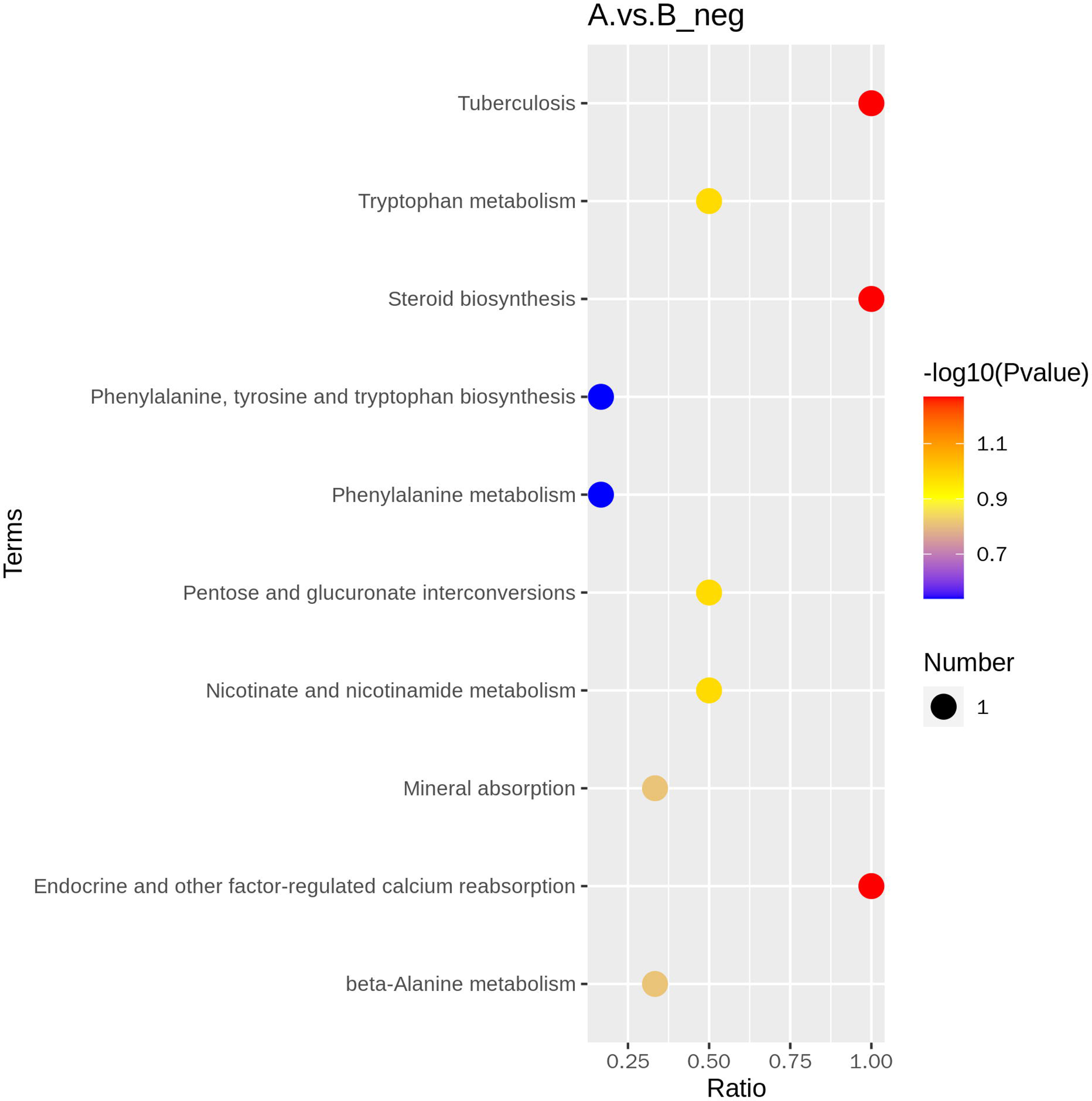
Significantly changed pathways based on the enrichment analysis. (A) KEGG enrichment bubble plot under the positive modes. (B) enrichment bubble plot under the positive modes.

### 3.7. Clinical correlations of selected metabolites

Spearman’s correlation analysis was employed to examine the relationship between metabolites and NT-proBNP. The 20 most significant metabolites identified through univariate regression demonstrated a moderate-to-high correlation with NT-proBNP (Figure 6). With the exception of PC 18:1_18:1 (r=-0.43, *p*=0.005),PC 18:1_20:5(r=-0.48, *p*=0.48), and LysoPE 18:2(r=-0.41, p=0.001), all other metabolites exhibited a predominantly positive correlation with NT-proBNP. This observation suggests that altered lipid metabolism, likely a consequence of metabolic stress, may play a critical role in the disease progression of HFpEF cases.

**Fig. 6.**
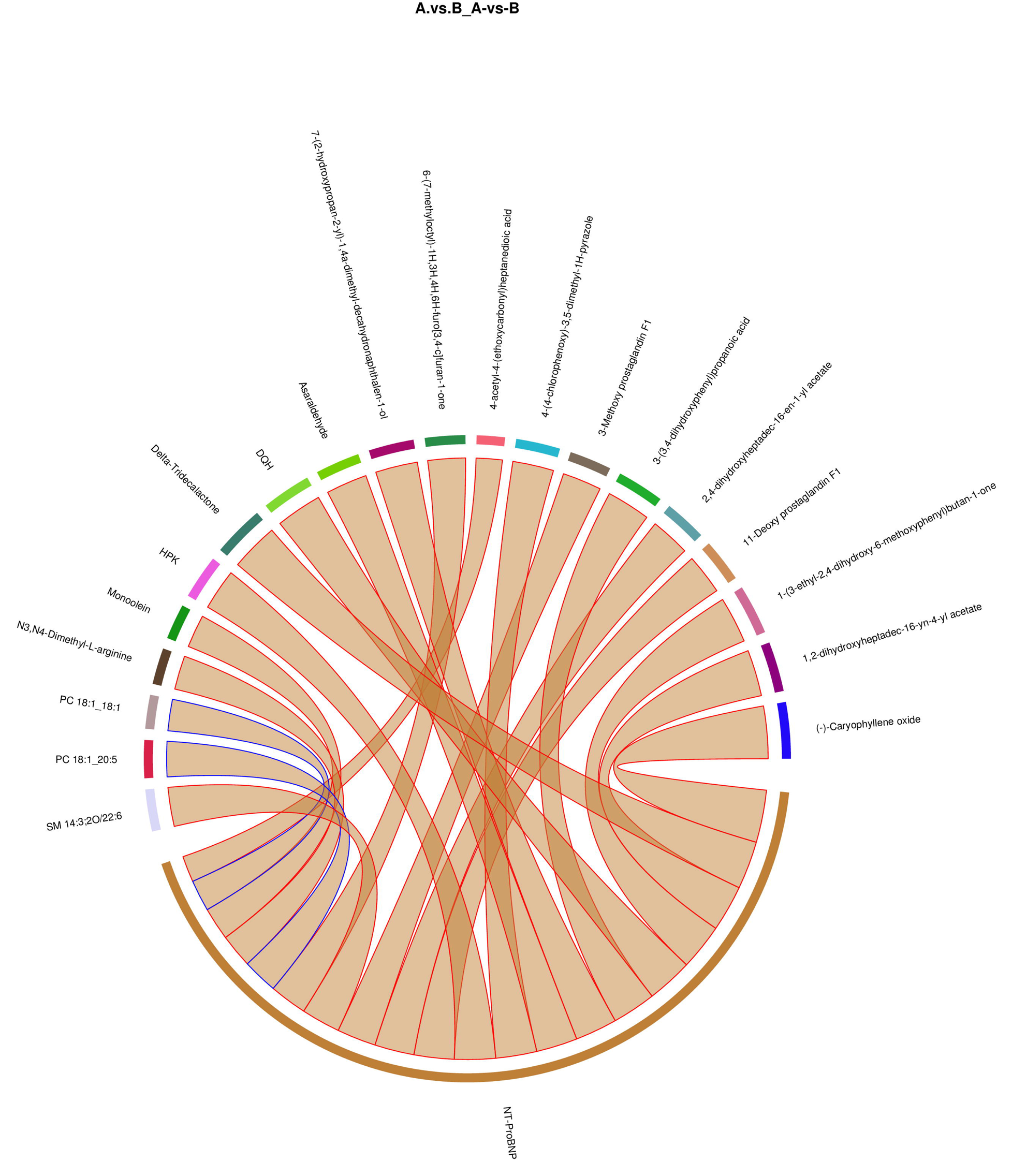
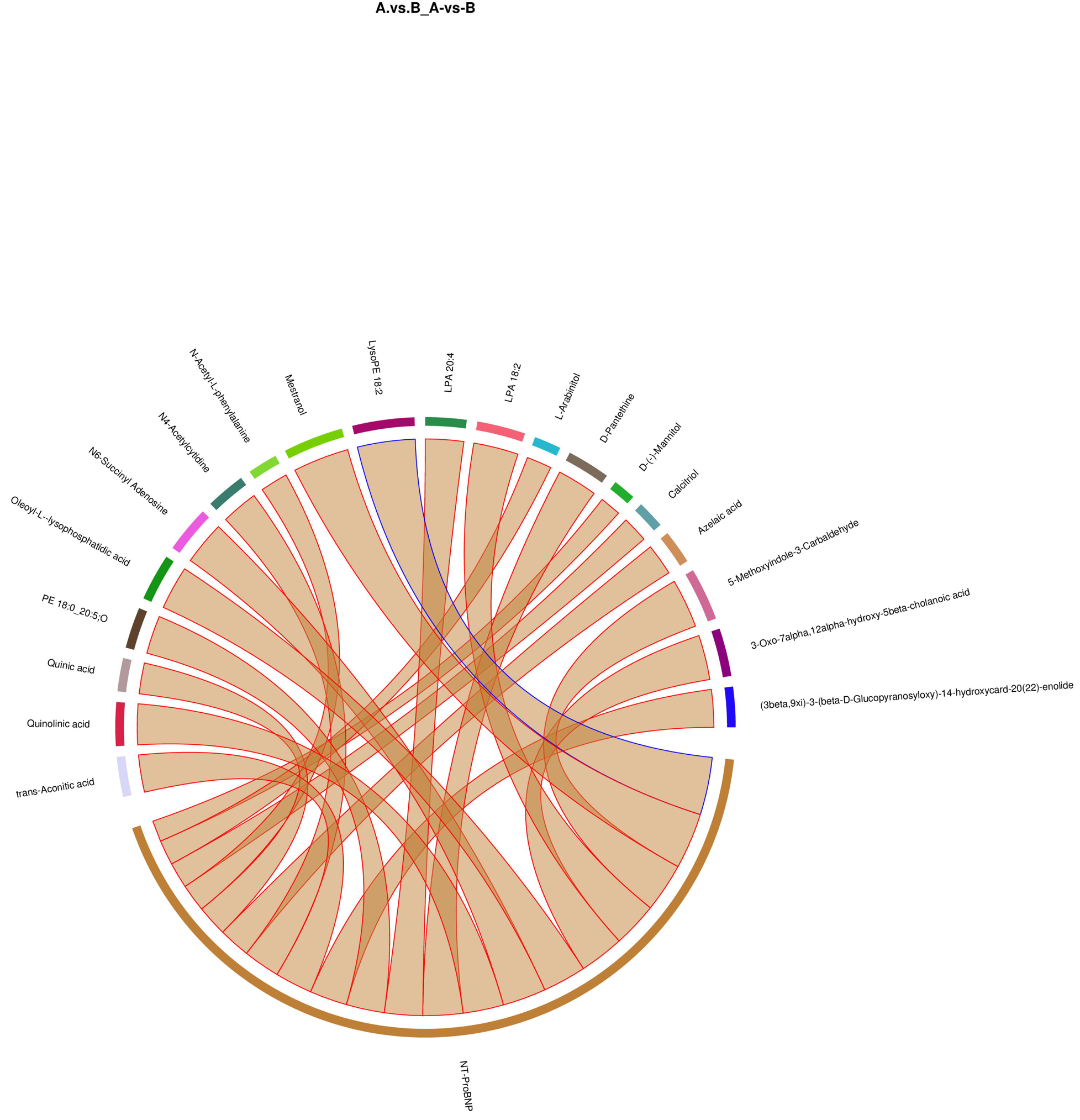
Correlation chord diagram (the upper the upper and lower frames show positive- and negative-ion mode, respectively). Note: Chord width represents correlation strength; Chord border color stands for correlation, with red and blue representing positive and negative correlations, respectively.

## 4. Discussion

Metabolic impairment significantly influences the onset and progression of HFpEF. However, the detailed metabolic pathogenesis of HFpEF remains largely unexplored. Previous investigations into the metabolic profiles of HFpEF have utilized various metabolomics approaches, including NMR spectroscopy, liquid chromatography-mass spectrometry (LC-MS) and gas chromatography-mass spectrometry (GC–MS) [10,16–19]. In this study, untargeted metabolomics testing identified differential metabolite expression profiles in the plasma of HFpEF patients and HC, highlighting distinct plasma metabolic characteristics in HFpEF compared to HC.

Initially, the most notable changes in HFpEF relative to HC were in amino acids, peptides, analogues, and lipids, followed by alterations in organoheterocyclic compounds. KEGG enrichment and pathway impact analysis indicated significant differences between the two groups. Notably, pathway such as Tryptophan metabolism, Tuberculosis and Endocrine and other factor-regulated calcium reabsorption were significantly altered, suggesting their potential critical impact on HFpEF progression. Additionally, 124 significantly different metabolites were selected, which Cytosine and 1,2-dihydroxyheptadec-16-yn-4-yl as potential biomarkers.

HFpEF is associated with oxidative stress and inflammation, impaired lipid metabolism, increased collagen production, disturbed lipid metabolism and reduced nitric oxide signaling [19]. Diabetes and obesity risk factors for HFpEF, contribute to left ventricular (LV) diastolic dysfunction, and cardiac lipotoxicity is believed to play a role in its pathogenic mechanism. Excessive fatty acids (FAs) are stored after consumption [20,21], and various lipids fact as signaling molecules in insulin resistance and inflammatory pathways [22,23], thereby influencing cardiovascular disease onset.

Glycerophospholipids, including PC, PE and lysophospholipids, are crucial in maintaining cell membrane structure and signal transduction. In HFrEF patients, serum levels of PC, lysoPC, lysoPE, and other substances significantly decrease, indicating a disruption in phospholipid metabolism linked to advanced age,poor clinical conditions, and impaired muscle oxidative metabolism [24]. Recent studies have also observed a significant decrease in serum levels of PC and lysoPC metabolites in the HFpEF patients [10,19]. our study, found notable decrease in glycerophospholipids(PC 18:1_20:5, PC 18:1_18:1, PC 36:2, PC O-40:8, PC 19:2_20:4, PC 20:3_20:4, PC 18:2_20:3, PC O-34:2, PC 19:2_19:2, PC 40:5, PC O-42:11, PC 19:2_18:5, PC O-36:2), PE(PE O-16:1_22:4, PE O-18:2_20:4) apparently decreased within plasma of HFpEF compared to HC group. these Glycerophospholipids play an important role in maintaining cell membrane integrity and signal transduction, and abnormal levels of these lipids can lead to myocardial metabolic disorders induced by lipotoxicity [25]. Furthermore, our study revealed a significant upregulation of phosphatidylcholines (PCs) and phosphatidylethanolamines (PEs), aligning with previous findings in HFpEF patients [10,19]. This supports the notion that disturbed lipid metabolism, due to metabolic stress is likely crucial for HFpEF progression.

Understanding the complexities of lipid metabolism in HFpEF could provide valuable insights for developing targeted treatments. Strategies focusing on modulating lipid uptake, enhancing lipid oxidation, and restoring mitochondrial function may help mitigate disturbances and improve cardiac function in HFpEF patients.

### Tryptophan metabolism

Tryptophan (Trp), an essential amino acid, serve as a precursor for various biochemical reactions in the human body, including the synthesis of serotonin, glycools, glucocorticoids and diabetic drugs [26]. Trp metabolism is closely associated with various cardiovascular diseases, with increasing research examining its relationship with Heart Failure(HF) [27–29].

Trp metabolism primarily involves pathways such as kynurenine(Kyn), 5-hydroxytryptamine, and indole, generating bioactive compounds that regulate functions like metabolism, inflammation, neurological function and immune responses [30]. The gut microbiota significantly affect Trp metabolism by transforming it into various molecules, including indole and its derivatives [31].

Recent studies link disturbances in Trp metabolism pathway and resultant Kyn up-regulation to myocardial infarction and atherosclerosis [32,33]. In this study, differential metabolites were highly enriched in the Trp metabolism pathway, particularly involving Indole-3-acetic acid and Kynurenic (Kyn) acid. these findings offer new insights into HFpEF’s pathological mechanisms and potential treatment strategies. energy metabolism, and signal transduction in myocardial cells [34]; Although research on Trp metabolism’s relationship with HFpEF is nascent, this discovery provides new insights into pathological mechanisms of HFpEF and the development of potential treatment strategies. For instance, regulating tryptophan metabolism could improve cardiac function and quality of life for HFpEF patients. However, several challenges remain, including unclear specific mechanisms by which Trp metabolism affects cardiac function, the lack of effective methods to regulate Trp metabolism, and the need for further research to determine the casual relationship and clinical significance of Trp metabolism in HFpEF.

In conclusion, the relationship between trp metabolism and HFpEF is an emerging research field with broad research prospects and clinical application potential. Future research needs to delve into the specific mechanisms of trp metabolism and how to utilize this mechanism to develop new treatment strategies.

## 5. Study Limitations

Certain limitations of the current study warrant mention. Firstly, the sample sizes or the HFpEF and HC groups were relatively modest, necessitating a larger, prospective validation cohort to substantiate the identified metabolites. Additionally, the metabolomic profile provides an overview of metabolic disturbances, potentially influenced by confounders such as acute illnesses, other disease states, and medication usage. Consequently, it remains uncertain whether the metabolic disturbances identified are exclusively associated with the HFpEF syndrome. Most current metabolomics studies, on heart failure patients utilize blood samples, However, Hahn et al. [1] recently reported that changes in plasma metabolite contents between HFpEF and HFrEF patients may not accurately reflect myocardial metabolic characteristics. This finding underscores the importance of using myocardial tissue directly for metabolomics analysis.

Overall, the small sample size in this study could impact the robustness of our results. the metabolome of each individual is highly sensitive to various endogenous and exogenous factors, such as age, sex, diet, environment, geographical location, genetics, and time of day [35].Therefore, future studies should focus on spontaneous screening of HFpEF patients and the validation of these findings. Current results could be further enriched and corroborated by integrating metabolomics research on plasma and faces.

## Data Availability

All data produced in the present study are available upon reasonable request to the authors

## Acknowledgement of grant support

This work was supported by the Key R&D Program of Xinjiang Uygur Autonomous Region [Grant No. 2022B03023-4].

